# Epidemiological profile of patients diagnosed with covid-19 in the municipality of São Gonçalo, Rio de Janeiro, Brazil

**DOI:** 10.1101/2024.09.08.24313185

**Authors:** Raphael Rangel das Chagas, Hércules Rezende Freitas, Sergian Vianna Cardozo

**Author notes:** Corresponding author: Raphael Rangel das Chagas, Interinstitutional Graduate Program in Translational Biomedicine (BIOTRANS), School of Health Sciences, Grande Rio University, Duque de Caxias, RJ, Brazil. Authors’ e-mail addresses: Raphael Rangel das Chagas, Hercules Rezende Freitas, Sergian Vianna Cardozo.

## Abstract

**Objectives:** The global health crisis caused by SARS-CoV-2 has led to over 760 million confirmed cases and 6.8 million deaths worldwide, primarily impacting the respiratory system with symptoms varying from mild to severe. This study aimed to analyze the interplay between vaccination status, sociodemographic profiles, comorbidities, and COVID-19 outcomes.

**Study Design:** Observational, cross-sectional, and analytical.

**Methods:** The study analyzed data from 6,953 individuals, examining vaccination statuses, sociodemographic profiles, comorbidities, COVID-19 test results, and other relevant variables. The cohort comprised predominantly mixed-race (51%), Caucasian (38%), and Black (9.5%) individuals, with 61% being female and 60% aged between 21-50 years. Prevalent comorbidities included hypertension (18.2%), diabetes (4.9%), and obesity (0.4%).

**Results:** Population-weighted analysis revealed significant associations between sociodemographic factors and COVID-19 test outcomes. Younger age groups, particularly 11-30 years, had higher positivity rates, which declined with age. Caucasians exhibited higher positivity rates (40.1%) compared to other ethnicities. Cramér’s V indicated small correlations between symptoms and test outcomes, notably with loss of taste (V = 0.11) and smell (V = 0.08). Odds ratio analysis identified hypertension as significantly associated with higher COVID-19 positivity (OR = 1.54, 95% CI: 1.28-1.83, p < 0.001), while obesity was associated with lower positivity (OR = 0.13, 95% CI: 0.02-0.63, p = 0.025). Symptoms such as fever, cough, loss of taste, loss of smell, and myalgia also showed significant associations with positive test outcomes.

**Conclusions:** This study provides valuable insights into the complex interplay of sociodemographic characteristics, comorbidities, symptoms, and COVID-19 outcomes.

## Introduction

SARS-CoV-2, a virus from the *Coronaviridae* family, *Betacoronavirus* genus, and *Sarbecovirus* subgenus, causes severe acute respiratory syndrome coronavirus 2 (SARS-CoV-2). This enveloped, roughly spherical virus measures 60-140 nm in diameter and has a single strand of positive-sense RNA, about 30,000 nucleotides long ^1^. SARS-CoV-2 is highly contagious, spreading through respiratory droplets and contact with contaminated surfaces ^2^.

The high transmissibility of SARS-CoV-2 led to an outbreak in Wuhan, China, in late 2019, sparking the global COVID-19 pandemic ^3^. The pandemic has become a significant public health challenge worldwide, resulting in over 760 million confirmed cases and 6.8 million deaths ^4^. The virus primarily affects the respiratory system, with symptoms ranging from mild to severe, including fever, cough, shortness of breath, fatigue, pneumonia, multiple organ failure, and death ^5^.

To combat the pandemic, various measures, including the development of vaccines, have been implemented. Vaccination is a primary strategy to prevent infection and reduce disease severity ^6,7^. The rapid development of COVID-19 vaccines has involved global efforts, with numerous candidate vaccines in progress^8–10^.

The pandemic has disrupted national vaccination schedules, particularly for children under 12 months, causing delays in their vaccinations ^11^. The continuous development of COVID-19 vaccines is crucial in controlling the virus spread and reducing cases.

In this study, we analyzed data from 6,953 individuals who received COVID-related care in São Gonçalo, Rio de Janeiro, Brazil. Our goal was to examine sociodemographic characteristics such as age, ethnicity, and gender in relation to COVID-19 testing results. We also aimed to identify common comorbidities, especially hypertension, among those who tested positive, and to investigate the prevalence of key symptoms, including fever above 38.5°C, loss of smell, and loss of taste, among participants who tested positive and negative.

## Methods

### Experimental design

The experimental design of this study is observational, cross-sectional, and analytical. The study population consists of patients who sought care at screening centers in São Gonçalo, a municipality in the state of Rio de Janeiro, Brazil, between July 28, 2021, and December 2, 2021.

According to the Brazilian Institute of Geography and Statistics (IBGE), São Gonçalo spans a territorial area of 248.160 km². In 2022, the resident population was estimated at 896,744 people, resulting in a population density of 3,613.57 inhabitants per km². The school enrollment rate for children aged 6 to 14 was 96.7% in 2010, and the Municipal Human Development Index (MHDI) was 0.739. Economically, the Gross Domestic Product (GDP) per capita was R$ 18,504.81 in 2021. Infant mortality, a critical health indicator, was 12.33 deaths per thousand live births in 2020 ^12^.

### Inclusion criteria and informed consent

Patients included in the study exhibited clinical symptoms suggestive of SARS-CoV-2 infection and underwent COVID-19 testing using the RT-LAMP technique. The screening centers in São Gonçalo served as locations for data collection and sample analysis.

This study adhered to the ethical principles outlined in the Declaration of Helsinki and relevant Brazilian legislation. Rigorous measures were implemented to ensure patient confidentiality, with all data anonymized before analysis. The research protocol was reviewed and approved by the Research Ethics Committee of the University of Grande Rio (CEP/UNIGRANRIO) under CAAE number 32362220.1.0000.5283.

All participants were thoroughly informed about the study’s objectives and potential risks and provided written informed consent to participate. Participation was voluntary, with no coercion or pressure applied, and participants retained the right to withdraw their consent at any time without facing any adverse consequences. The study was conducted following the highest ethical standards, ensuring the integrity and dignity of both participants and researchers.

### Data collection

Data collection at screening centers in São Gonçalo, Rio de Janeiro, included sociodemographic characteristics, comorbidities, COVID-19 symptoms, vaccination history, and COVID-19 test results. Patient age was determined by recording the date of birth and the date of sample collection. Ethnicity was documented to understand ethnic diversity. Comorbidities such as hypertension, diabetes, and obesity were detailed, along with other comorbidities.

The onset date of COVID-19 symptoms and specific symptoms reported, including runny nose, fever, loss of smell and taste, diarrhea, headache, shortness of breath, muscle pain, and cough, were recorded. COVID-19 test results, using the RT-LAMP technique, were documented as positive or negative. The number of COVID-19 vaccine doses received, the type of vaccine administered, and the date of administration were also recorded.

Healthcare professionals, adhering to safety and ethical protocols, conducted data collection. Participants were informed about the data collection purpose and provided written informed consent. Data confidentiality was maintained, and information was handled anonymously.

### Statistical analysis

The statistical analysis for this study was conducted using RStudio (version 2023.06.1 + 524) with the R programming language (version 4.2.2).

Through univariate analysis, we aimed to describe the profile of the participants, identify the number of non-responses, and detect outliers. Data presented in Tables 1 to 6 include the analysis of sociodemographic characteristics, comorbidities, COVID-19 test results, and other relevant variables. This analysis was conducted using absolute and relative frequencies as well as mean values.

**Table 1.** Sociodemographic characteristics of the cohort of participants (patients) who were attended at the screening centers of São Gonçalo (RJ), during the period from 07/28/2021 to 12/02/2021.

| Variables | N = 6,953<sup>1</sup> |
| --- | --- |
| AGE |  |
| ≤ 10 y.o. | 10 (0.1%) |
| 11 to 20 y.o. | 491 (7.1%) |
| 21 a 30 y.o. | 1,419 (20%) |
| 31 a 40 y.o. | 1,406 (20%) |
| 41 a 50 y.o. | 1,457 (21%) |
| 51 a 60 y.o. | 1,142 (16%) |
| 61 a 70 y.o. | 679 (9.8%) |
| 71 a 80 y.o. | 274 (3.9%) |
| ≥ 81 y.o. | 75 (1.1%) |
| ETHNICITY |  |
| Brown | 3,490 (51%) |
| Caucasian | 2,625 (38%) |
| Black | 652 (9.5%) |
| Other | 88 (1.3%) |
| Not available | 98 |
| SEX |  |
| Female | 4,263 (61%) |
| Male | 2,690 (39%) |
| <sup>1</sup>n (%) |  |

Bivariate analysis was performed to examine significant associations between study variables. Considerring that the data was collected from a convenience sample, weights were calculated by post-stratification based on the known populational (IBGE, 2024) ethnicity and sex distributions. Then, the Pearson’s chi-square test (χ²) with Rao & Scott adjustment was employed to evaluate the associations between study variables:

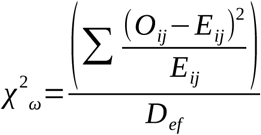

Where *χ²_ω_*represents the (weighted) adjusted chi-square test statistic, *Oij* denotes the observed frequency counts, *Eij* denotes the expected frequency counts, and *D_ef_* is the calculated design effect. This methodological approach is consistent with the literature on statistical data analysis in health, as highlighted by ^13^, who emphasize the importance of univariate and bivariate analyses and the application of the Pearson chi-square test for categorical variable associations.

Additionally, to assess the strength of associations identified by the *χ²* test, a weighted Cramer’s V was calculated:

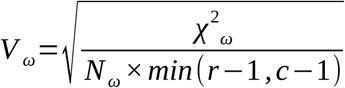

Where *V_ω_*indicates the strength of association between variables (Cramer’s V), *χ²_ω_*is the weighted *χ²* test statistic, N *_ω_* is the total weight (sum of all weights), *r* is the number of rows in the contingency table, and *c* is the number of columns in the contingency table. For interpretive purposes, *χ²_ω_*and *V_ω_* values with *p* less than 0.05 were considered statistically significant, allowing for the rejection of the null hypothesis.

Finally, to analyze the relationship between various study variables, odds ratios (OR) were calculated, providing a measure of association between exposure variables and the outcome variable, which in this case is the result of the COVID-19 test. The OR was calculated as:

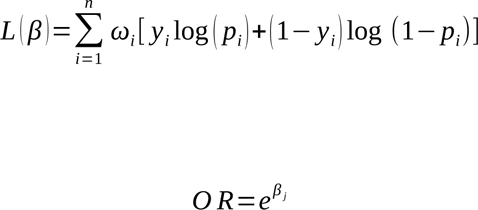

Where *L(b)* is the log-likelihood function, *p_i_* is the predicted probability that *Y_i_* = 1, *OR* is the weighted odds ratio for a given predictor, and *b_j_* is the estimated coefficient for the predictor. For each calculated OR, a 95% confidence interval (CI) was also computed to assess the precision of the estimate. Also, for interpretative purposes, a p-value less than 0.05 considered statistically significant. This approach ensures that the identified associations are less likely to derive from random chance.

## Results

Table 1 details the sociodemographic characteristics of the study participants, categorizing them by age group, ethnicity, and gender. Age groups ranged from 10 to 81 years and above. Ethnicity was classified into mixed-race (parda), Caucasian, Black, other, and unknown. The data indicate that most participants were mixed-race (51%), followed by Caucasian (38%) and Black (9.5%). Furthermore, a significant proportion of the participants were female (61%). Age-wise, the largest group was aged 21 to 50 years, comprising approximately 60% of the participants. Those aged 61 to 80 years constituted around 14%, while participants aged 81 years or older made up about 1% of the total sample.

Key comorbidities (*i.e.*, obesity, hypertension, and type II diabetes) were also examined (Supplementary table 1). Most participants did not report having diabetes (95.1%), obesity (99.6%), or hypertension (81.8%). Among the comorbidities reported, hypertension was the most prevalent (18.2%), followed by diabetes (4.9%) and obesity (0.4%). This highlights hypertension as the most common comorbidity within the study population.

Table 2 presents additional variables beyond the comorbidities and sociodemographic characteristics of the study participants. It details the number of participants who experienced symptoms such as fever, runny nose, loss of smell, loss of taste, diarrhea, headache, shortness of breath, and muscle pain. The table also provides information on the participants’ vaccination status, including the number of doses received and the type of vaccine administered.

**Table 2.** Symptoms, immunization, and geographical distribution in the sample of participants (patients) who were attended at the screening centers of São Gonçalo (RJ), during the period from 07/28/2021 to 12/02/2021.

| Variables | N = 6,953<sup>1</sup> |
| --- | --- |
| FEVER < 38.5 °C |  |
| No | 5,057 (73%) |
| Yes | 1,828 (27%) |
| Not available | 68 |
| FEVER ≥ 38.5 °C |  |
| No | 5,893 (86%) |
| Yes | 989 (14%) |
| Not available | 71 |
| RHINITIS |  |
| No | 4,496 (65%) |
| Yes | 2,382 (35%) |
| Not available | 75 |
| LOSS OF SMELL |  |
| No | 5,908 (86%) |
| Yes | 956 (14%) |
| Not available | 89 |
| LOSS OF TASTE |  |
| No | 5,912 (86%) |
| Yes | 968 (14%) |
| Not available | 73 |
| DIARRHEA |  |
| No | 6,354 (92%) |
| Yes | 544 (7.9%) |
| Not available | 55 |
| HEADACHE |  |
| No | 3,483 (51%) |
| Yes | 3,414 (49%) |
| Not available | 56 |
| DYSPNEA |  |
| No | 6,052 (88%) |
| Yes | 824 (12%) |
| Not available | 77 |
| MYALGIA |  |
| No | 4,556 (66%) |
| Yes | 2,316 (34%) |
| Not available | 81 |
| COUGH |  |
| No | 3,268 (48%) |
| Yes | 3,580 (52%) |
| Not available | 105 |
| RESULT |  |
| Negative | 4,925 (72%) |
| Positive | 1,953 (28%) |
| Not available | 75 |
| DOSE |  |
| Unvaccinated | 53 (1.0%) |
| First dose | 1,578 (30%) |
| Second dose | 3,298 (62%) |
| Third dose | 206 (3.9%) |
| Single dose | 142 (2.7%) |
| Not available | 1,676 |
| VACCINE |  |
| Oxford - AstraZeneca | 2,352 (45%) |
| Coronavac - Sinovac | 1,163 (22%) |
| Pfizer - BNT162b2 | 1,532 (29%) |
| Janssen-Cilag | 149 (2.9%) |
| Not available | 1,757 |
| MUNICIPALITY |  |
| São Gonçalo | 6,851 (99%) |
| Other | 102 (1.5%) |
| VACCINATED |  |
| No | 53 (1.0%) |
| Yes | 5,224 (99%) |
| Not available | 1,676 |

<sup>1</sup>n (%)

The data indicate that most participants did not have a fever above 38.5°C (73.4%), runny nose (65.6%), loss of smell (86%), or loss of taste (85.9%). Moreover, the majority received two vaccine doses (62.5%), with the Oxford-AstraZeneca vaccine being the most administered (45%). Among the variables examined, headache was the most frequently reported symptom, affecting 49% of the participants.

Table 3 shows the relationship between the sociodemographic profile of participants and their COVID-19 test results, detailing age, ethnicity, and gender. Younger individuals, specifically those aged 11-20 years, showed a higher positivity rate (7.4%) compared to the negative group (6.8%). This trend continued with the 21-30 years (18.0% positive) and 31-40 years (17.8% positive) groups, though these groups also had substantial negative test rates (21.3% and 21.1%, respectively). Positivity rates decreased with age, with a notable difference in the 71-80 years group (6.8% positive vs. 2.8% negative). The 81+ years group had 1.3% positive and 1.0% negative.

**Table 3.** Relationship between the sociodemographic profile and the COVID-19 test results of participants (patients) who were attended at the screening centers of São Gonçalo (RJ), during the period from 07/28/2021 to 12/02/2021.

| Variables | COVID-19 TEST RESULT |  |  | p-value<sup>2</sup> |
| --- | --- | --- | --- | --- |
|  | TOTAL, N =<br>6,878<sup>1</sup> | NEGATIVE, N =<br>4,925<sup>1</sup> | POSITIVE, N =<br>1,953<sup>1</sup> |  |
| AGE |  |  |  | 0.000* |
| ≤ 10 y.o. | 10 (0.1%) | 6 (0.1%) | 4 (0.2%) |  |
| 11 to 20 y.o. | 482 (7.0%) | 337 (6.8%) | 145 (7.4%) |  |
| 21 a 30 y.o. | 1,399 (20.3%) | 1,048 (21.3%) | 351 (18.0%) |  |
| 31 a 40 y.o. | 1,386 (20.2%) | 1,039 (21.1%) | 347 (17.8%) |  |
| 41 a 50 y.o. | 1,441 (21.0%) | 1,063 (21.6%) | 378 (19.4%) |  |
| 51 a 60 y.o. | 1,137 (16.5%) | 796 (16.2%) | 341 (17.5%) |  |
| 61 a 70 y.o. | 676 (9.8%) | 448 (9.1%) | 228 (11.7%) |  |
| 71 a 80 y.o. | 272 (4.0%) | 139 (2.8%) | 133 (6.8%) |  |
| ≥ 81 y.o. | 75 (1.1%) | 49 (1.0%) | 26 (1.3%) |  |
| ETHNICITY |  |  |  | 0.002* |
| Brown | 3,446 (50.8%) | 2,490 (51.2%) | 956 (49.9%) |  |
| Caucasian | 2,605 (38.4%) | 1,836 (37.7%) | 769 (40.1%) |  |
| Black | 646 (9.5%) | 478 (9.8%) | 168 (8.8%) |  |
| Other | 84 (1.2%) | 61 (1.3%) | 23 (1.2%) |  |
| Not available | 97 | 60 | 37 |  |
| SEX |  |  |  | 0.394 |
| Female | 4,226 (61.4%) | 3,096 (62.9%) | 1,130 (57.9%) |  |
| Male | 2,652 (38.6%) | 1,829 (37.1%) | 823 (42.1%) |  |

<sup>1</sup>n (%)
<sup>2</sup>Pearson $\chi^2$ test

Ethnicity showed significant variations. The brown ethnic group had nearly half (49.9%) test positive, slightly lower than the negative group (51.2%). Caucasians had a higher positivity rate (40.1%) compared to their negative counterparts (37.7%). The black ethnic group had a slightly lower positivity rate (8.8%) compared to the negative group (9.8%). Other ethnicities had almost identical positive (1.2%) and negative (1.3%) rates. A small portion lacked ethnic classification, which did not significantly impact overall trends.

Gender analysis revealed no significant differences. Females constituted 57.9% of positive cases and 62.9% of negative cases. Males represented 42.1% of positive cases and 37.1% of negative cases.

Table 4 illustrates the relationship between comorbidities and COVID-19 test results. It details the number of participants who reported having hypertension, diabetes, and obesity, as well as the number of participants who tested positive or negative for COVID-19 within each comorbidity group.

**Table 4.**
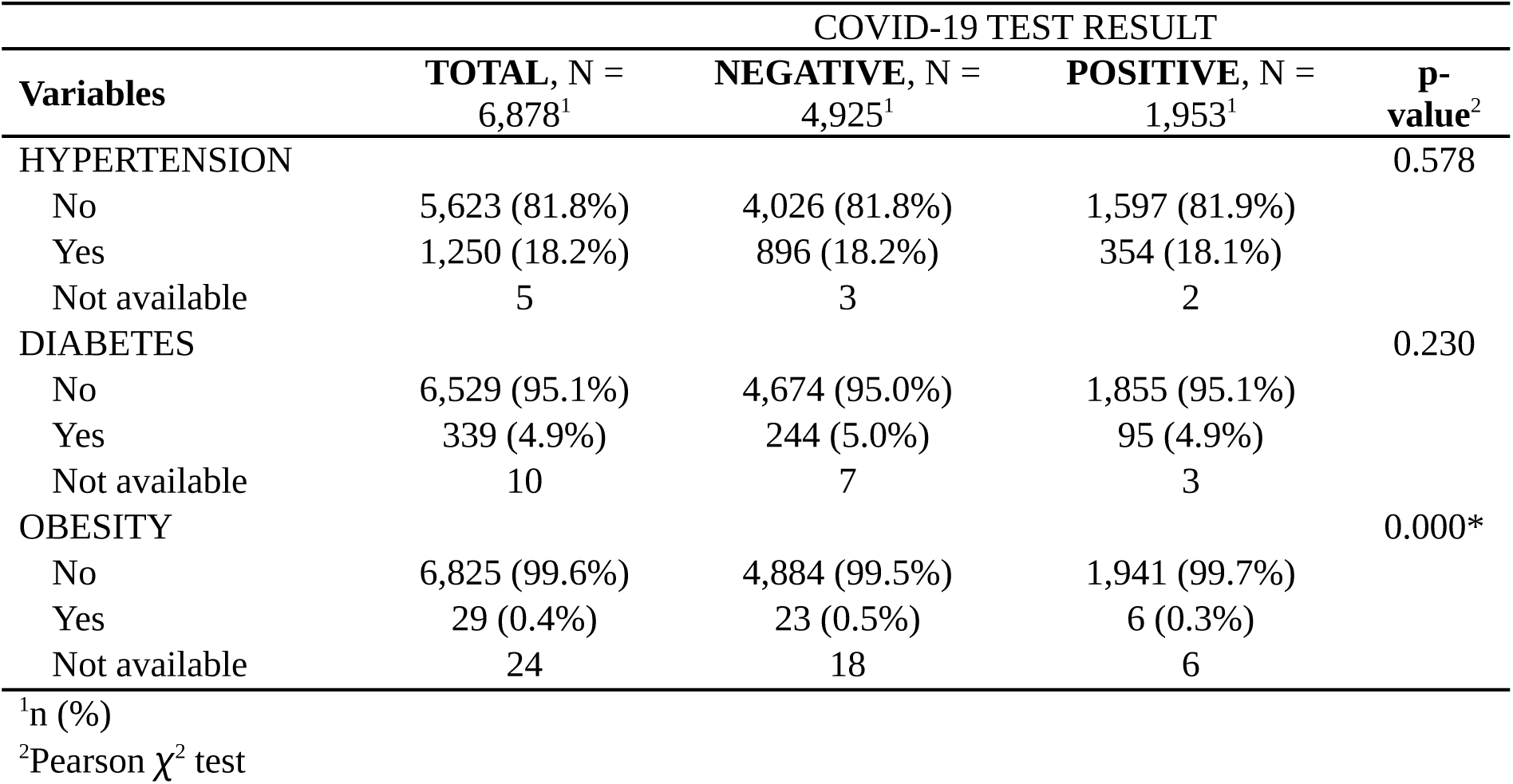
Comorbidities and COVID-19 test results.

| Variables | COVID-19 TEST RESULT |  |  | p-value<sup>2</sup> |
| --- | --- | --- | --- | --- |
|  | TOTAL, N = 6,878<sup>1</sup> | NEGATIVE, N = 4,925<sup>1</sup> | POSITIVE, N = 1,953<sup>1</sup> |  |
| HYPERTENSION |  |  |  | 0.578 |
| No | 5,623 (81.8%) | 4,026 (81.8%) | 1,597 (81.9%) |  |
| Yes | 1,250 (18.2%) | 896 (18.2%) | 354 (18.1%) |  |
| Not available | 5 | 3 | 2 |  |
| DIABETES |  |  |  | 0.230 |
| No | 6,529 (95.1%) | 4,674 (95.0%) | 1,855 (95.1%) |  |
| Yes | 339 (4.9%) | 244 (5.0%) | 95 (4.9%) |  |
| Not available | 10 | 7 | 3 |  |
| OBESITY |  |  |  | 0.000* |
| No | 6,825 (99.6%) | 4,884 (99.5%) | 1,941 (99.7%) |  |
| Yes | 29 (0.4%) | 23 (0.5%) | 6 (0.3%) |  |
| Not available | 24 | 18 | 6 |  |

<sup>1</sup>n (%)<sup>2</sup>Pearson $\chi^2$ test

The analysis of comorbidities and their association with COVID-19 test results provides several key insights. Hypertension did not show a significant link to COVID-19 test outcomes. Among those without hypertension, the positive test rate was 81.9%, nearly identical to the negative test rate of 81.8%. Similarly, 18.1% of individuals with hypertension tested positive, compared to 18.2% who tested negative.

Diabetes also did not exhibit a significant correlation with COVID-19 test results. The data showed that 95.1% of individuals without diabetes tested positive, while 95.0% tested negative. Among those with diabetes, 4.9% tested positive compared to 5.0% negative. In contrast, obesity showed a significant association with COVID-19 test results. The overwhelming majority of individuals without obesity tested positive (99.7%) and negative (99.5%). However, among those with obesity, only 0.3% tested positive compared to 0.5% negative.

Table 5 shows the relationship between symptoms, vaccination status, and COVID-19 test results. It categorizes participants based on the presence of fever >38.5°C, runny nose, loss of smell, loss of taste, diarrhea, headache, shortness of breath, muscle pain, and cough, as well as their vaccination status, and compares the number of positive and negative test results.

**Table 5.**
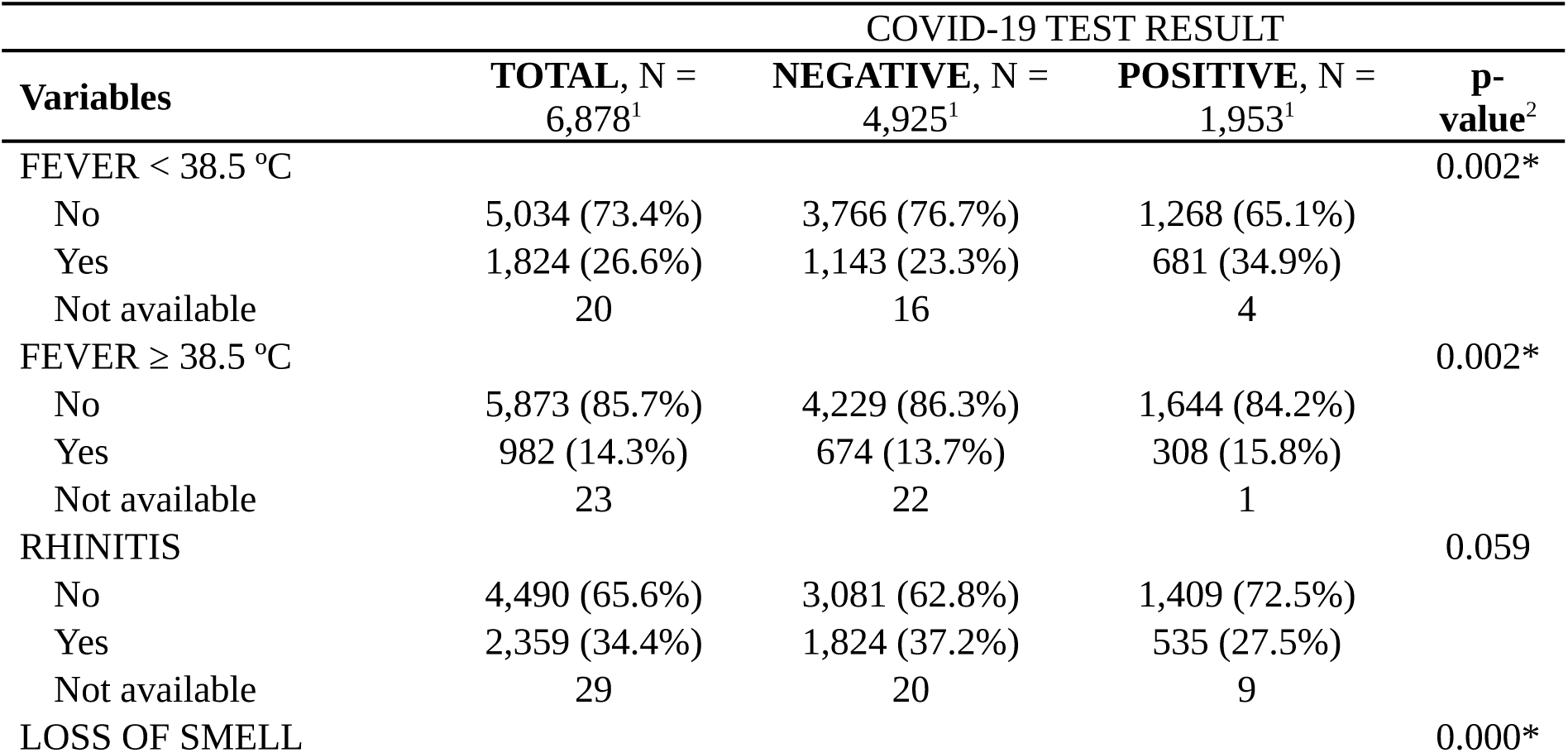
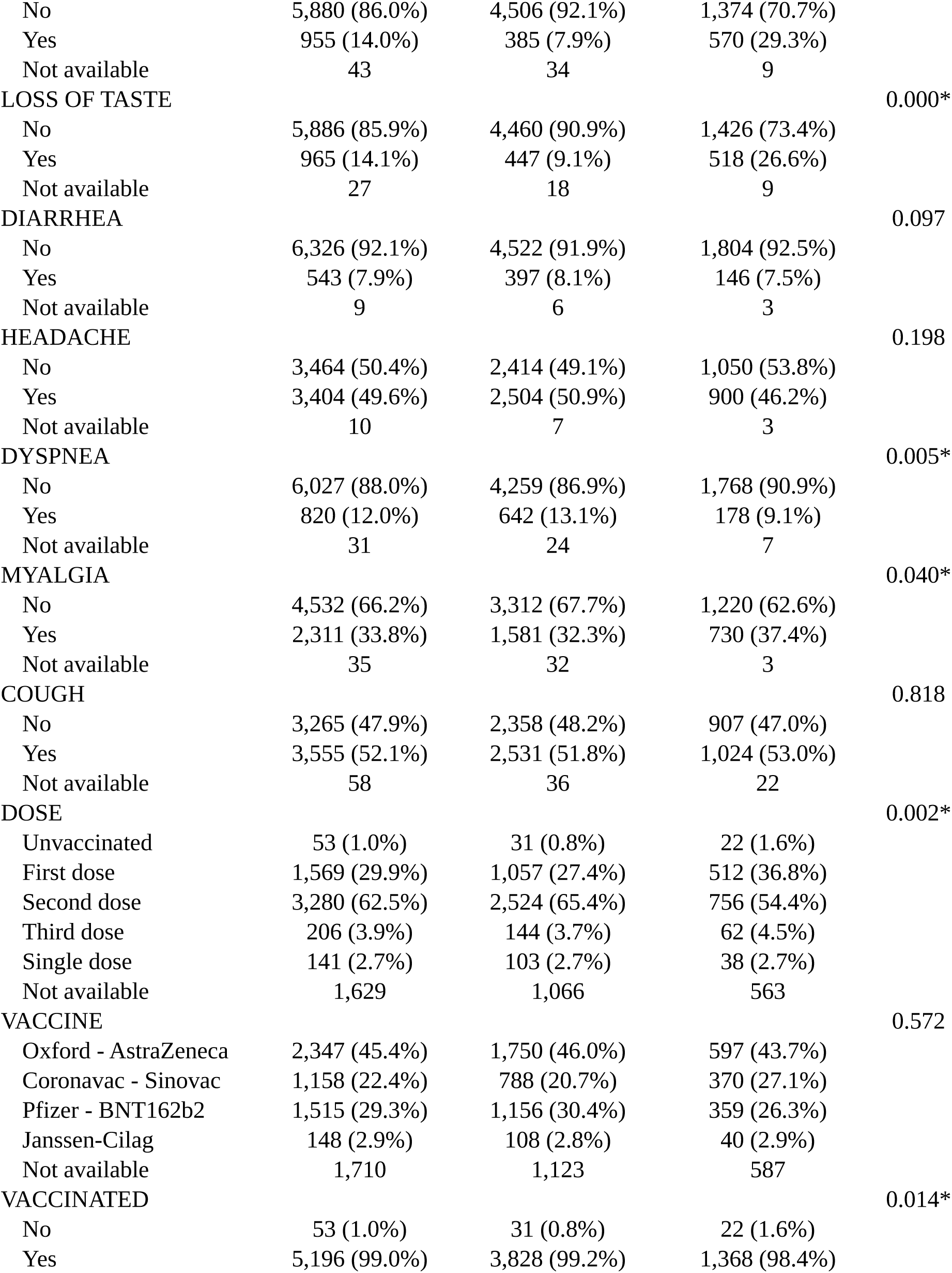
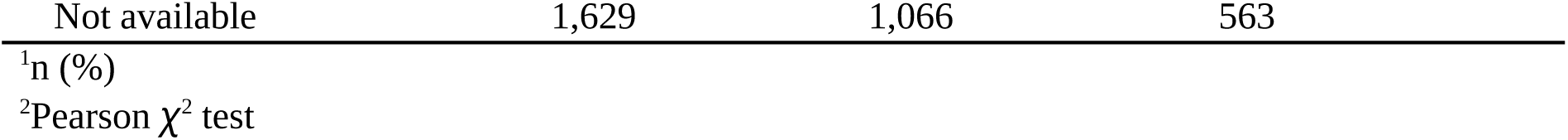
Other variables and the COVID-19 test results of participants (patients) who were attended at the screening centers of São Gonçalo (RJ), during the period from 07/28/2021 to 12/02/2021.

A significant proportion of positive cases had fever >38.5°C (15.8%), fever ≤38.5°C (34.9%), loss of smell (29.3%), and loss of taste (26.6%). Most negative cases did not have these symptoms. Dyspnea was more common in negative cases (13.1% vs. 9.1%), while myalgia was more frequent in positive cases (37.4% vs. 32.3%).

Regarding vaccination, 36.8% of first-dose recipients tested positive, while 27.4% tested negative. For second-dose recipients, 54.4% tested positive, and 65.4% tested negative. Third-dose recipients had a positive test rate of 4.5%, slightly higher than the negative rate of 3.7%. Single-dose recipients had an identical positive and negative rate of 2.7%.

The type of vaccine did not show a significant association with test results. Among Oxford-AstraZeneca recipients, 43.7% tested positive, and 46.0% tested negative. CoronaVac-Sinovac had a positive rate of 27.1% and a negative rate of 20.7%. Pfizer-BNT162b2 had a positive rate of 26.3% and a negative rate of 30.4%. Janssen-Cilag had a positive rate of 2.9% and a negative rate of 2.8%. This suggests comparable effectiveness across different vaccines.

Overall vaccination status significantly related to test results. Among the unvaccinated, 1.6% tested positive, while 0.8% tested negative.

In addition to analyzing the independence of the study variables, the strength of the associations between the variables of interest and parameters such as ethnicity, vaccination status, and the number of vaccine doses received was evaluated using the same sample.

As depicted in Figure 1, loss of taste and smell show the greater (although weak) correlations with the test results (Cramér’s V = 0.08 and 0.11, respectively). Low and high fever (highly intercorrelated, V = 0.25), as well as dyspnea, showed equaly negligible (although significant) correlations with test results (Cramér’s V = 0.04). Finally, myalgia was the symptom with the smaller significant correlation with test results (Cramér’s V = 0.03).

**Figure 1.**
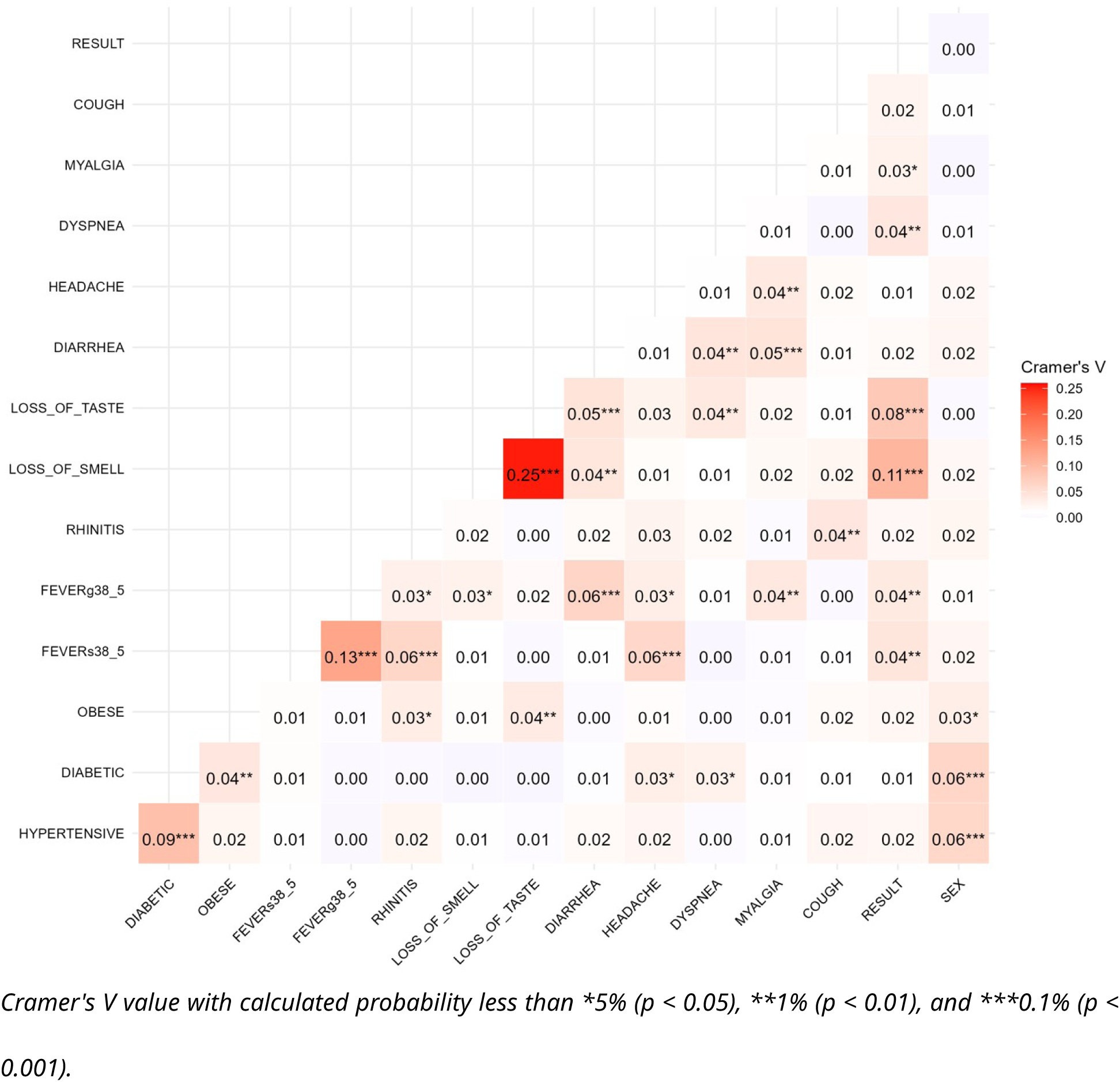
Heatmap of the magnitude of association (weighted Cramer’s V) between the study variables and vaccination parameters, ethnicity, and vaccine dose.

Here, we also calculated odds ratios for various study variables against COVID-19 test results, shown in Figure 2. Hypertension was significantly associated with higher likelihood of testing positive (OR = 1.54, 95% CI: 1.28-1.83; p = 2.67^-6^). Diabetes showed a trend towards higher likelihood but was not significant (OR = 1.30, 95% CI: 0.97-1.73; p = 0.078). Obesity was associated with a significantly lower likelihood (OR = 0.13, 95% CI: 0.02-0.63; p = 0.025). Fever >38.5°C had a strong association (OR = 2.55, 95% CI: 2.17-3.00; p = 9.26^-30^), and fever ≤38.5°C also showed significant association (OR = 2.11, 95% CI: 1.80-2.48; p = 6.68^-20^). Rhinitis was linked to lower likelihood (OR = 0.68, 95% CI: 0.59-0.78; p = 3.94^-8^). Loss of smell (OR = 3.65, 95% CI: 2.91-4.59; p = 8.48^-29^) and loss of taste (OR = 2.05, 95% CI: 1.62-2.57; p = 1.14 ^-9^) were strong indicators of positive tests.

**Figure 2.**
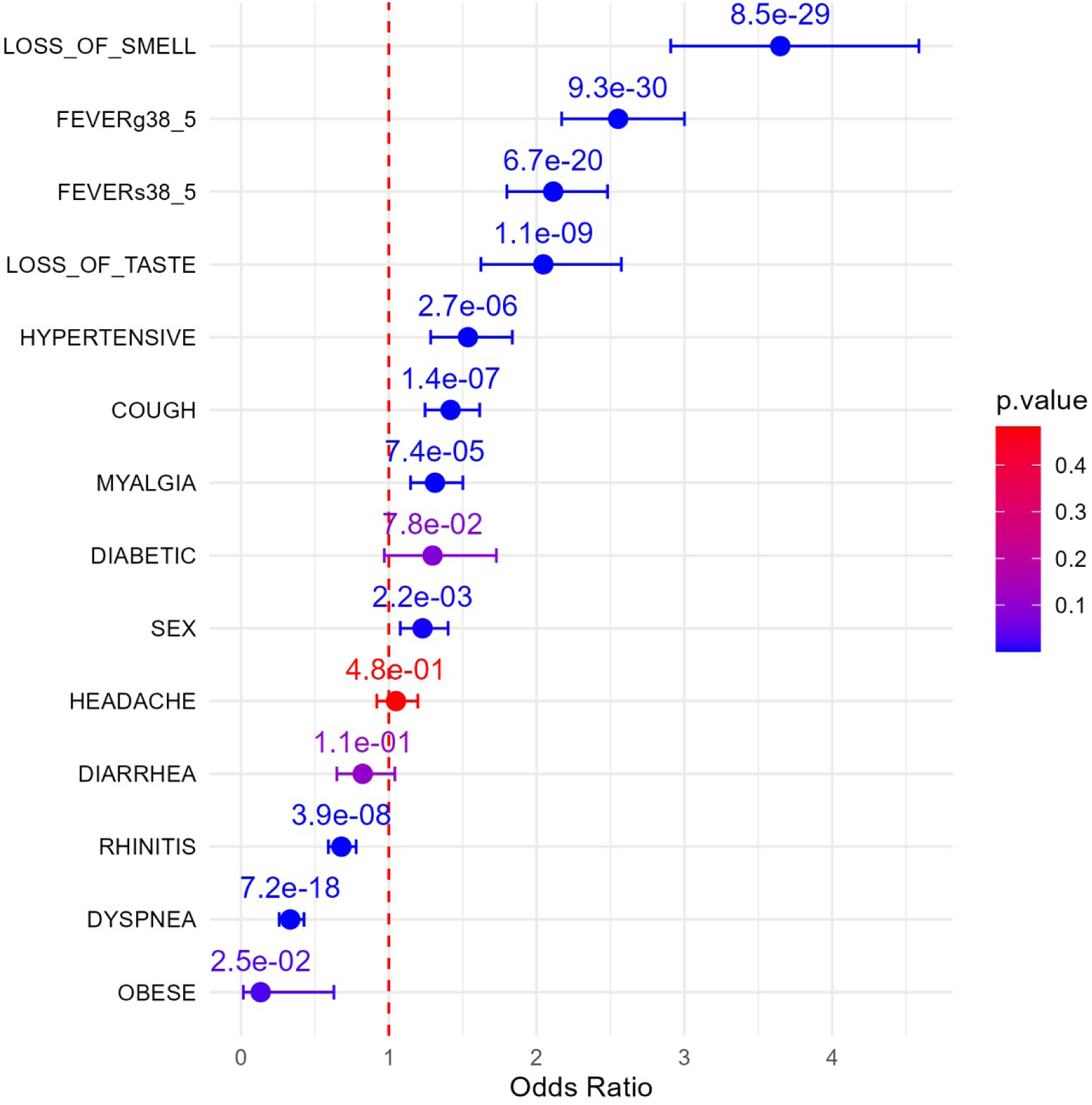
Weighted odds ratios evaluating study variables against covid-19 test results.

Diarrhea showed a non-significant trend towards lower likelihood (OR = 0.82, 95% CI: 0.65-1.04; p = 0.107). Headache showed no significant association (OR = 1.05, 95% CI: 0.92-1.20; p = 0.483). Dyspnea was strongly associated with lower likelihood (OR = 0.33, 95% CI: 0.26-0.43; p = 7.21^-18^). Myalgia (OR = 1.31, 95% CI: 1.15-1.50; p = 7.38^-5^) and cough (OR = 1.42, 95% CI: 1.25-1.62; p = 1.43^-7^) were significantly associated with higher likelihood. Being male was also associated with higher likelihood (OR = 1.23, 95% CI: 1.08-1.40; p = 0.002).

## Discussion

### Sociodemographic Variables

Age, race, ethnicity, and gender significantly impact COVID-19 infection and outcomes. According to Table 4, 74.5% of individuals who tested positive for COVID-19 were over 31 years old, with the highest prevalence among those aged 41 to 50 years (19.4%). Age significantly influences COVID-19 status and progression, with individuals over 65 exhibiting elevated viral loads, higher pro-inflammatory markers, and reduced immune response efficiency due to factors like inflammation, immunological aging, and changes in ACE2 receptor dynamics and cytokine profiles^14^. The mental state of the elderly can also impact their recovery ^15^.

Racial and ethnic minority groups have been disproportionately affected by the pandemic ^16^. In the current cohort, 59.9% of those testing positive for COVID-19 identified as non-white (mixed race, Black, and other ethnicities). African Americans, Hispanics, and non-Hispanic Blacks have higher infection rates than other racial and ethnic groups ^17,18^. Socioeconomic status and healthcare access significantly contribute to these disparities. Lower to middle socioeconomic status is linked to higher COVID-19 hospitalization rates ^19^. Non-white participants face increased exposure risks and healthcare barriers, leading to higher chances of seroconversion and hospitalization ^20^. These findings highlight the need for equitable resource distribution and targeted interventions to address COVID-19’s disproportionate impact on racial and ethnic minority groups.

Data from ten European countries indicate that working-age women have higher COVID-19 diagnosis rates than men, with the trend reversing after retirement. Women’s higher infection rates may be due to their greater representation in healthcare-related occupations ^21^. In the current study, 57.9% of individuals testing positive for COVID-19 were female. The relationship between gender and COVID-19 infection is complex and multifaceted. Research shows gender disparities in vulnerability, severity, and outcomes of COVID-19. Despite men being more susceptible to severe infection outcomes, including higher hospitalization and mortality rates compared to women ^22^, women, due to genetic and hormonal factors, exhibit a more resilient immune response. Women’s expression of the TLR7 gene on the X chromosome enhances their ability to produce antiviral proteins and interferons in dendritic cells, protecting them against severe SARS-CoV-2 infections ^22^. Additionally, cardiovascular conditions significantly affect COVID-19 outcomes, with men exhibiting greater vulnerability compared to women^23^.

### Comorbidities

Hypertension is a primary risk factor for adverse COVID-19 outcomes, associated with severe illness and increased mortality. In ^24^, 74% of hypertensive patients with moderate symptoms died, with a relative risk (RR) of 1.45 (CI 0.96-2.2). ^25^ reported a hazard ratio (HR) of 1.58 (CI 1.07-2.32) linked to hypertension, and any comorbidity presence was associated with an HR of 1.79 (CI 1.16-2.77). Another study found a 22% odds ratio (OR) (1.22; CI 1.12-1.33) increase in severe infection among hypertensive patients. Both hypertensive (>180 mm/Hg, OR 1.93; CI 1.06-3.51) and hypotensive (<120 mm/Hg, OR 1.40; CI 1.11-1.78) patients had higher chances of severe COVID-19 ^26^.

Hypertension may increase the risk of contracting COVID-19. For instance, in a study of 140 hospitalized COVID-19 patients, 30% had hypertension ^27^. Another study with 1099 confirmed COVID-19 patients reported hypertension in 23.7% ^28^. However, these studies do not establish a causal relationship between hypertension and COVID-19 infection. Conversely, other studies found no significant association between hypertension and COVID-19 infection risk. A retrospective study of 1178 hospitalized COVID-19 patients in Wuhan found no difference in hypertension prevalence (30.7%) between severe and non-severe cases or survivors and non-survivors, suggesting hypertension and ACE inhibitor use may not significantly increase infection risk ^29^.

Diabetes is strongly associated with severe symptoms, worse outcomes, and higher mortality in COVID-19 patients. Patients with diabetes often have poorer prognoses and increased susceptibility to infections due to complications like peripheral neuropathy and vascular insufficiency, nearly doubling the risk of death ^30–33^. Recent-onset hyperglycemia and new-onset diabetes have been reported in COVID-19 patients without pre-existing diabetes, suggesting a bidirectional interaction ^34–36^. However, whether people with diabetes are inherently more prone to COVID-19 infection remains unclear. Overall, diabetes is consistently associated with worse outcomes and increased mortality.

Obesity is also linked to severe COVID-19 outcomes and mortality ^37^. Studies show obese patients are more likely to develop severe disease and require invasive mechanical ventilation ^38,39^. Obesity reduces cardiorespiratory reserves and exacerbates immune dysfunction, increasing risks of hospitalization, respiratory infections, airway diseases, fatty liver, hypertension, depression, type 2 diabetes, cardiovascular diseases, and cancer ^40^. An extensive review identified obesity as a risk factor for severe COVID-19, with patients having BMI >40 kg/m^2^ showing higher odds for mechanical ventilation (2.17; CI 1.59-2.97) and mortality (1.67; CI 1.39-2.00)^41^.

In the present study, hypertension was the most common comorbidity (18.1% of infected patients), followed by diabetes (4.9%) and obesity (0.3%). Only obesity frequencies differed significantly between negative and positive patients (0.5% vs. 0.3%, *χ*^2^ p = 0.000).

### Symptomatology

Fever, anosmia, ageusia, cough, fatigue, shortness of breath, myalgia/arthralgia, chills, nausea/vomiting, nasal congestion, diarrhea, abdominal pain, hemoptysis, and conjunctival congestion are common COVID-19 symptoms ^42,43^. Gastrointestinal symptoms like nonspecific abdominal pain, watery diarrhea, and ileus have also been reported ^44^. Pulmonary symptoms include pneumonia, ARDS, and respiratory failure, while arrhythmias and neurological symptoms are extrapulmonary manifestations ^45^. Long-term symptoms include lethargy, difficulty concentrating, and cough. Prolonged symptoms one month post-infection are associated with being female, having chronic lung disease, and hypertension ^46^.

COVID-19 requires differential diagnosis due to nonspecific symptoms ^47^. It includes other respiratory diseases like influenza and pre-existing lung pathologies ^48^. Symptoms such as cough, fever, shortness of breath, and headache overlap with other respiratory infections and flu-like illnesses ^49^. Radiological features of COVID-19 pneumonia, like ground-glass opacities, crazy-paving patterns, and consolidations, challenge differential diagnosis ^50^. A multidisciplinary approach and specific diagnostic tests, such as SARS-CoV-2 antigen determination, are crucial for accurate diagnosis.

In the present study, the most prevalent symptoms in positive patients were high fever (≥ 38.5°C; 15.8%, p = 0.002), low fever (< 38.5°C; 34.9%, p = 0.002), loss of smell (29.3%, p = 0.000), loss of taste (26.6%, p = 0.000), and myalgia (37.4%, p < 0.04). Testing positive for COVID-19 is linked to symptoms like fever (both high and low), loss of smell, loss of taste, and myalgia, as documented in the literature ^51^. Myalgia, common in viral disorders and other inflammatory conditions, often presents in patients with hypertension and fever, indicating a systemic response to infections ^52^. Changes in taste and smell, distinctive of COVID-19 ^53^, are also associated with metabolic conditions like obesity, where systemic inflammation can affect sensory function ^54^. High fever, a marker of systemic inflammatory response, can signal complications in patients with hypertension or diabetes, where inadequate control of underlying conditions may predispose to complications ^55^.

### Vaccination

Table 5 highlights a significant relationship between the number of vaccine doses and negative COVID-19 test outcomes (p = 0.002). COVID-19 vaccination status (vaccinated or not vaccinated) was significantly linked to test results (negative or positive) (p = 0.014, Pearson’s *χ*^2^ test). These findings underscore that vaccination significantly reduces the risk of testing positive for COVID-19, with most positive cases occurring in unvaccinated individuals, while the majority of vaccinated individuals test negative.

Independent evidence supports this relationship. Studies have indicated a higher proportion of COVID-19 positive cases among unvaccinated individuals compared to those who have received at least one vaccine dose ^56^. Additionally, vaccination has been shown to reduce risks of hospitalization, ICU admissions, and oxygen requirement ^57^. Vaccination plays a pivotal role in mitigating severe COVID-19 cases, especially in fully vaccinated individuals. Social determinants such as age, income, education, and migration history also influence COVID-19 vaccination uptake, with higher rates among older adults, individuals with higher education and income levels, and lower rates among those with a migration history ^58^. Recent clinical trials have underscored the efficacy of mRNA vaccines in preventing virus infection ^59^. These insights highlight the critical role of vaccination in preventing COVID-19 infection and reducing disease severity.

## Final remarks

Our data offers a comprehensive view of sociodemographic characteristics, comorbidities, symptoms, and COVID-19 test outcomes among participants. Key findings include:

- Participants were predominantly of mixed race, followed by Caucasian and Black individuals, with females outnumbering males. The age group 21-50 years was most represented, while those 81 years or older were least represented.
- Most participants did not report hypertension, diabetes, or obesity, although hypertension was the most prevalent among those who did.
- Reported symptoms varied widely, with headache being the most common. Fever above 38.5°C, loss of smell, and loss of taste were frequent among those testing positive for COVID-19, but a significant proportion of positive cases did not show these symptoms.
- Age significantly affected the likelihood of testing positive.
- Hypertension was more commonly associated with positive COVID-19 tests among participants with comorbidities, while diabetes and obesity were less prevalent among those testing positive.
- Fever, loss of smell, loss of taste, and headache were key indicators of infection. Vaccination significantly reduced positive test rates, with a higher proportion of unvaccinated participants testing positive.

In conclusion, these insights can guide targeted strategies for prevention, screening, and treatment, and identify at-risk populations.

## Data Availability

All data produced in the present study are available upon reasonable request to the authors

## Acknowledgements

Ethical approval

The present research protocol was reviewed and approved by the Research Ethics Committee of the University of Grande Rio (CEP/UNIGRANRIO) under CAAE number 32362220.1.0000.5283.

## Supplementary tables

**Supplementary table 1.** Comorbidities present in the sample of participants (patients) who were attended at the screening centers of São Gonçalo (RJ), during the period from 07/28/2021 to 12/02/2021.

| Variables | N = 6,953<sup>1</sup> |
| --- | --- |
| HYPERTENSION |  |
| No | 5,652 (82%) |
| Yes | 1,253 (18%) |
| Not available | 48 |
| DIABETES |  |
| No | 6,560 (95%) |
| Yes | 339 (4.9%) |
| Not available | 54 |
| OBESITY |  |
| No | 6,855 (100%) |
| Yes | 31 (0.5%) |
| Not available | 67 |

<sup>1</sup>n (%)

